# HAVAs: Alzheimer’s Disease Detection using Normative and Pathological Lifespan Models

**DOI:** 10.1101/2021.12.03.21267230

**Authors:** Pierrick Coupé, José V. Manjón, Boris Mansencal, Thomas Tourdias, Gwenaëlle Catheline, Vincent Planche

## Abstract

In this paper, we present an innovative MRI-based method for Alzheimer’s Disease (AD) detection and mild cognitive impairment (MCI) prognostic, using lifespan trajectories of brain structures. After a full screening of the most discriminant structures between AD and normal aging based on MRI volumetric analysis of 3032 subjects, we propose a novel Hippocampal-Amygdalo-Ventricular Alzheimer score (HAVAs) based on normative lifespan models and AD lifespan models. During a validation on three external datasets on 1039 subjects, our approach showed very accurate detection (AUC ≥ 94%) of patients with AD compared to control subjects and accurate discrimination (AUC=78%) between progressive MCI and stable MCI (during a 3 years follow-up). Compared to normative modelling and recent state-of-the-art deep learning methods, our method demonstrated better classification performance. Moreover, HAVAs simplicity makes it fully understandable and thus well-suited for clinical practice or future pharmaceutical trials.

## 1 Introduction

Finding early and specific biomarkers of Alzheimer’s disease (AD) clinical syndrome is of major interest to accelerate the development of new therapies. Among the potential structural biomarkers proposed for AD, neurodegeneration estimated using magnetic resonance imaging (MRI) is still a good candidate [1], [2]. From simple volume-based approaches to advanced deep learning strategies, the development of new biomarkers able to detect anatomical alterations caused by AD has been the subject of much attention over the past decades [3]–[5].

Nowadays, two main strategies are used to detect neurodegeneration caused by AD using MRI: normative modelling for abnormality detection [6], [7] and classification-based approaches [8], [9].

On the one hand, normative modelling based only on cognitively normal (CN) subjects can be used to detect abnormality and therefore to distinguish AD patients from CN subjects. As explained in [7], normative lifespan modelling is similar to growth charts used in pediatric medicine to detect abnormal child development in terms of height or weight related to the age’s subject. Indeed, such charts can be used to detect outliers considered as pathological. For AD detection, volume or thickness of key structures as a function of age is usually used. The main advantages of normative modelling are to robustly capture the heterogeneity of normal anatomy and to provide an easily interpretable distance between an individual and the normative range. Normative modelling is the approach used in most of the available software for quantitative brain analysis (in open access such as volBrain [10] or for commercial use as in Neuroquant^®^ [11], Qscore^®^ [12] or Qreport^®^ [13]). The added-value in terms of diagnosis accuracy has been shown for several pathologies including AD [11]–[14]. Due to its simplicity and easy understanding, normative modelling is the closest strategy to clinical practice with several CE marked and FDA approved software packages.

On the other hand, a classifier can be trained using features extracted from the two groups – one composed of CN subjects and another one composed of AD patients. The used features can be handcrafted as usually done in Machine Learning (ML) [3] or automatically learned using Deep Learning (DL) [15]. At the end of the training, a decision boundary is available to discriminate features of CN subjects from features of AD patients. Such a strategy is supposed to be more accurate than normative modelling since patients are used in addition to CN subjects during training. Consequently, the developed method is pathology specific. Moreover, by using advanced methods such as DL, a specific signature of a given pathology can be automatically and efficiently learned. However, such approaches suffers from a lack of generalization usually related to overfitting on the training database [8], [16]. Moreover, with the advent of DL methods, interpretation of the results and explanation of the underlying decision-making process is far from being straightforward [15].

In this paper, we present an alternative framework combining advantages of both strategies: an easy interpretation and an accurate classification. To this end, we propose a novel method able to detect patients with AD using both normal and pathological lifespan models. First introduced in [17], lifespan modelling of AD provides an useful and easily interpretable tool to capture the heterogeneity of AD signature. Moreover, by using multiple models (i.e., an AD model in addition to a CN model), the decision boundary is pathology specific and thus produces a more accurate detection of AD patients compared to usual normative modelling. Finally, we also propose an innovative framework to extract the most discriminant structures between both groups based on a fully automatic multi-scale brain segmentation pipeline. Applied to AD, this framework led us to propose a novel Hippocampal-Amygdalo-Ventricular Alzheimer score (HAVAs) based on multiple lifespan models.

## 2 Material and Method

### 2.1 Dataset description

#### 2.1.1 Training dataset

Our training dataset was composed of 3032 T1-weighted (T1w) MRI from seven open access databases (see Table 1). This dataset was composed of 2655 CN subjects (CN) and 377 patients with AD. As explained in the following, CN subjects younger than 55y (N= 1874) were used to estimate both CN and AD lifespan trajectories.

**Table 1:** Training dataset description used for model constructions after quality control (N=3032). This table provides the name of the databases, the group, the number of considered subjects, the gender proportion, and the average age with the interval in brackets.

| DATASET | Group | N=3032 | Gender | Age in years |
| --- | --- | --- | --- | --- |
| C-MIND | CN | 236 | F = 129 / M = 107 | 8.44 [0.74-18.86] |
| NDAR | CN | 382 | F = 174 / M = 208 | 12.39 [1.08-49.92] |
| ABIDE | CN | 492 | F = 84 / M = 408 | 17.53 [6.50-52.20] |
| ICBM | CN | 294 | F = 142 / M = 152 | 33.75 [18-80] |
| IXI | CN | 549 | F = 307 / M = 242 | 48.76 [20.0- 86.2] |
| OASIS | CN | 298 | F = 187 / M = 111 | 45.34 [18 - 94] |
| ADNI | CN | 404 | F = 203 / M = 201 | 74.81 [60 – 90] |
| OASIS | AD | 45 | F = 29 / M = 16 | 77.04 [63 - 96] |
| ADNI | AD | 332 | F = 151 / M = 181 | 75.13 [55 – 91] |

#### 2.1.2 Testing dataset

To validate our model, we built a testing dataset based on two open access databases (AIBL and MIRIAD) to perform AD vs. CN diagnosis task. Therefore, we validated the generalization capacity of our method and its robustness to domain shift. In addition, we used subjects with Mild Cognitive Impairment (MCI) from ADNI to estimate the capability of our models on prognosis task (see Table 2). Consequently, we validated the generalization of our models to unseen related tasks. As in [8], the MCI group was split into stable MCI (sMCI) over three years and progressive MCI (pMCI) who will convert to AD within 36 months following the baseline visit.

**Table 2:** External dataset used for validation (N=1039). This table provides the name of the databases, the group, the number of considered subjects, the gender proportion, and the average age with the interval in brackets.

| DATASET | Group | N=1039 | Gender | Age in years |
| --- | --- | --- | --- | --- |
| AIBL | CN | 467 | F = 277 / M = 190 | 73.4 [60.5 – 92.4] |
| MIRIAD | CN | 23 | F = 11 / M = 12 | 69.7 [58.0 – 85.7] |
| ADNI | sMCI | 255 | F = 100 / M = 155 | 72.3 [55 – 89.5] |
| AIBL | AD | 82 | F = 47 / M = 36 | 74.8 [55.5 – 93.4] |
| MIRIAD | AD | 46 | F = 27 / M = 19 | 69.3 [55.6 – 85.8] |
| ADNI | pMCI | 235 | F = 103 / M = 132 | 74.0 [55 – 88.0] |

### 2.2 Image processing

All the considered images were processed using AssemblyNet software^1^ [18]. Based on collective artificial intelligence, AssemblyNet is able to produce fine-grained segmentation of the whole brain in 15 minutes. The AssemblyNet preprocessing pipeline was based on several steps: image denoising [19], inhomogeneity correction [20], affine registration to the MNI space, automatic quality control (QC) [21], a second inhomogeneity correction in the MNI space [22] and a final intensity standardization step [10].

After preprocessing, the brain was segmented into several structures using 250 DL models (see [18] for details). All the segmentations were based on the Neuromorphometrics protocol which comprises 132 structures [23]. In this protocol, the segmentation of the subcortical structures follows the “general segmentation protocol” as defined by the MGH Center for Morphometric Analysis^2^. Moreover, the segmentation of the cortical structures follows the “BrainCOLOR protocol”^3^. These structures are combined to create tissue segmentations (gray matter (GM), white matter (WM) and cerebrospinal fluid (CSF)), regional tissue segmentations (cortical GM, subcortical GM, ventricular CSF and external CSF) and lobar segmentations (temporal, limbic, insular, parietal and frontal) – see Figure 1.

**Figure 1:**
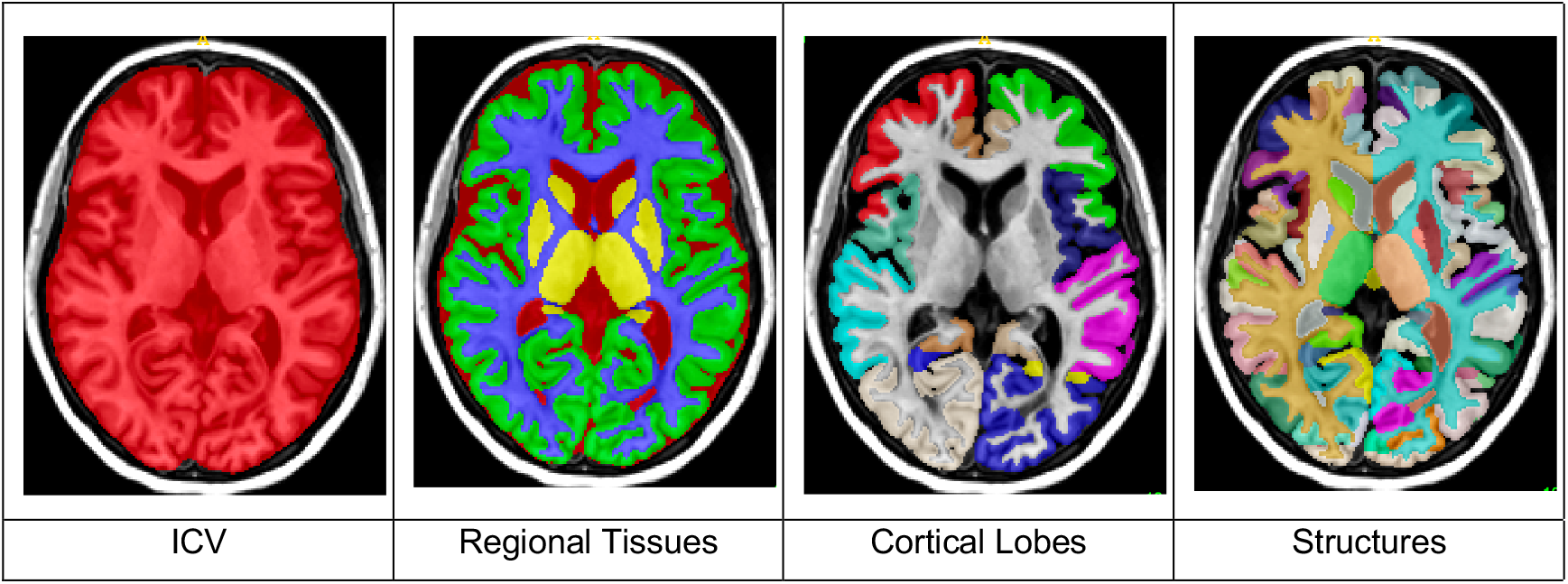
Illustrations of the AssemblyNet multi-scale segmentations.

Finally, we performed a QC procedure to carefully select subjects included in our training dataset. For all the training subjects detected as failure by the automatic QC RegQCNet [21], a visual assessment was performed by individually checking the input images and the segmentations produced by AssemblyNet using a 3D viewer. If the failure was confirmed by our expert, the subject was removed from training dataset.

### 2.3 Volume normalization

To compensate for the inter-subject variability, we normalized all the structure volumes using the intracranial cavity volume (ICV) [24]. Moreover, in order to be able to combine several structures with different sizes, we performed z-score normalization of all the normalized volumes (in percentage of ICV). To do that, we first estimated the mean and the standard deviation for each structures using all the CN subjects over the entire lifespan. Then, for a given structures, we applied the same z-score normalization to all the subjects (i.e., CN, AD and MCI). Therefore, by using z-score of normalized volumes in % of ICV, we compensated for both inter-subject and inter-structure variabilities. In the following, all the volumes are expressed as z-scores of normalized volumes.

### 2.4 Lifespan model estimation

To create our lifespan models, we estimated normal and pathological trajectories of structure volumes across the entire lifespan. To this end, for each considered structure, models were estimated on two different groups to generate CN and AD trajectories. For CN trajectories, we used the N=2655 subjects from 9 months to 94y of the training dataset as done in [25]. For the AD trajectories, we used N=2251 subjects. As done in [17], we mixed AD patients with young CN. More precisely, we used 377 AD patients (from 55y to 96y) and all the CN younger than 55y available in the training dataset (i.e., 1874 subjects) assuming that neurodegeneration is a slow and progressive process.

To estimate the volume trajectories, we considered several low order polynomial models:

- Linear model

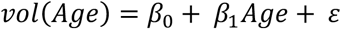
- Quadratic model

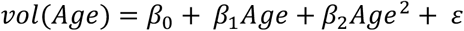
- Cubic model

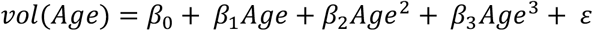

As in [17], [25], a polynomial model was considered as a potential candidate only when simultaneously F-statistic based on ANOVA (i.e., model vs. constant model) was found significant (p<0.05) and when all its coefficients were also significant using T-statistic (p<0.05). Afterwards, to select the most relevant model between these potential candidates, we used the Bayesian Information Criterion [26]. In addition, we estimated the distance between both AD and CN models as the Euclidean distance between trajectories. Finally, we estimated the confidence interval for each model at 95% and the lifetime period for which the two models diverged significantly (i.e., when confidence intervals do not overlap).

### 2.5 Classification using volume trajectories

Once the AD and CN lifespan trajectories were estimated for each structure using the training dataset, we used them to perform subject classification. For each subject of the testing dataset, we estimated the closest trajectory to assign the class of the subject under study. Moreover, we estimated scores of being an AD patient or a CN subject based on the distance to the models. To define these scores, we used the following approach.

First, for GM and WM structures, we defined a score *s*_*CN*_ to be CN (respectively *s*_*AD*_ to be AD) based on the distance to CN model (respectively to AD model) taking into account structure atrophy:

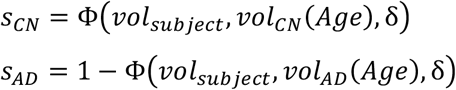

Where Φ(z, μ, σ) is the cumulative distribution function of the standard normal distribution of mean μ and standard deviation σ. In our case, we used δ = |*vol*_*CN*_(*Age*) *− vol*_*AD*_(*Age*)| to take into account the increasing distance between the both models during aging.

For CSF structures, we adapted the estimation taking into account structure enlargement caused by AD [27] as follows:

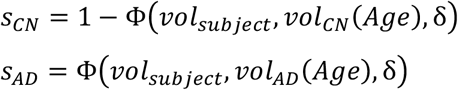

Finally, these scores were normalized to obtain the final scores. This normalization enables to get the sum of both scores equal to 1.

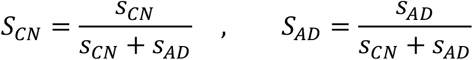

The classification performance of the proposed method was validated using several metrics: balanced accuracy (BACC), specificity (SPE), sensibility (SEN) and Area Under the Curve (AUC).

### 2.6 Comparison with normative and DL methods

Finally, we compared the proposed multi-model HAVAs with both normative model-based strategy (i.e., using only CN model) and state-of-the-art deep learning methods. As usually done in normative modelling [7] or in automatic quantitative software [13], we used 2σ as threshold to detect abnormal values when using normative model-based methods. We decided to evaluate lifespan normative approach using hippocampus (considered as the state-of-the-art biomarker [1]), amygdala (also known to be a good candidate [17]), inferior lateral ventricle (main part of lateral ventricle impacted by AD [28]) and the combination of the three as done for the proposed HAVAs (called Normative HAV model in the following).

As shown in [8], most of the proposed deep learning methods suffer from data leakage resulting in biased reported performances. Consequently, we used the well-evaluated methods proposed in [8] as state-of-the-art deep learning methods. We selected a ROI-based Convolutional Neural Network (CNN) focused on hippocampal area, one subject-based CNN method using the entire image and one patch-based CNN processing the whole image patch by patch. These three strategies are a good representation of current deep learning frameworks for AD detection and prognosis.

## 3 Results

### 3.1 Detection of the most discriminant structures

First, we selected all the multi-scale brain areas (i.e., tissues, regional tissues, lobes and structures) for which CN and AD models significantly diverged (i.e., confidence intervals stop overlapping at some point across lifespan). Thanks to this analysis, we obtained 33 areas. Using these 33 selected areas, we performed a screening to detect the most discriminant ones in terms of classification accuracy on the training ADNI dataset in order not to use testing data during method development. This analysis showed that amygdala, hippocampus and inferior lateral ventricle were the most discriminant structures for AD vs. CN classification (see Table 3). These three structures obtained AUC>80% and thus were selected to build our AD-specific hybrid lifespan models.

**Table 3:** Performance of the classification using multiple lifespan models on the training ADNI dataset (404 CN vs. 332 AD) for the 33 selected structures. The best results are indicated in red and second best in green. Finally, “n.s.” means that the divergence of frontal lobe was not significant.

|  | BACC | SPE | SEN | AUC |
| --- | --- | --- | --- | --- |
| WM | 61 | 53 | 69 | 69 |
| CSF | 66 | 60 | 71 | 73 |
| - External CSF | 59 | 53 | 64 | 64 |
| - Ventricular CSF | 68 | 72 | 64 | 71 |
| ▪ Inf. Lat. Vent | 75 | **85** | 64 | 82 |
| ▪ Lat. Vent | 68 | 70 | 65 | 71 |
| GM | 66 | 64 | 68 | 70 |
| - Subcortical GM | 70 | 66 | 73 | 75 |
| ▪ Amygdala | **82** | **85** | **79** | **88** |
| ▪ Hippocampus | **80** | **78** | **81** | **87** |
| ▪ Accumbens Area | 59 | 52 | 66 | 64 |
| ▪ Putamen | 57 | 53 | 60 | 61 |
| ▪ Thalamus | 56 | 55 | 58 | 62 |
| ▪ Pallidum | 55 | 55 | 55 | 58 |
| ▪ Caudate | 57 | 52 | 62 | 61 |
| - Cortical GM | 61 | 59 | 63 | 69 |
| ○ Temporal lobe | 71 | 71 | 71 | 78 |
| ▪ Middle temporal gyrus | 66 | 66 | 66 | 63 |
| ▪ Fusiform gyrus | 63 | 61 | 66 | 72 |
| ▪ Inferior temporal gyrus | 62 | 60 | 64 | 68 |
| ▪ Superior temporal gyrus | 60 | 59 | 62 | 65 |
| ▪ Temporal pole | 61 | 60 | 63 | 67 |
| ○ Limbic cortex | 64 | 61 | 67 | 68 |
| ▪ Entorhinal area | 64 | 64 | 63 | 71 |
| ▪ Parahippocampal gyrus | 64 | 65 | 63 | 70 |
| ▪ Anterior cingulate gyrus | 59 | 54 | 64 | 63 |
| ○ Insular cortex | 60 | 57 | 63 | 63 |
| ▪ Anterior insula | 58 | 55 | 61 | 63 |
| ▪ Posterior insula | 58 | 56 | 59 | 63 |
| ○ Parietal lobe | 57 | 53 | 60 | 59 |
| ▪ Angular gyrus | 59 | 55 | 64 | 63 |
| ○ Frontal lobe | *n.s.* | *n.s.* | *n.s.* | *n.s.* |
| ▪ Middle frontal gyrus | 55 | 52 | 57 | 58 |

### 3.2 Combination of the main AD MRI-based biomarkers

Based on our screening, we decided to combine the volume of hippocampus, amygdala and inferior lateral ventricle to propose a novel Hippocampal-Amygdalo-Ventricular Alzheimer score (HAVAs). To do that, we simply added hippocampus and amygdala volumes and subtracted the inferior lateral ventricle volume. Indeed, contrary to hippocampus and amygdala showing lower volumes in AD model due to atrophy, inferior lateral ventricle exhibited larger volumes in AD model due to enlargement. As done before, HAVAs is also expressed as a z-score of normalized volume. As shown in Figure 2, HAVAs exhibited an earlier divergence between CN and AD models (i.e., it can be used on younger subjects) and a larger distance between models (i.e., it is more discriminant) compared to single structure models.

**Figure 2:**
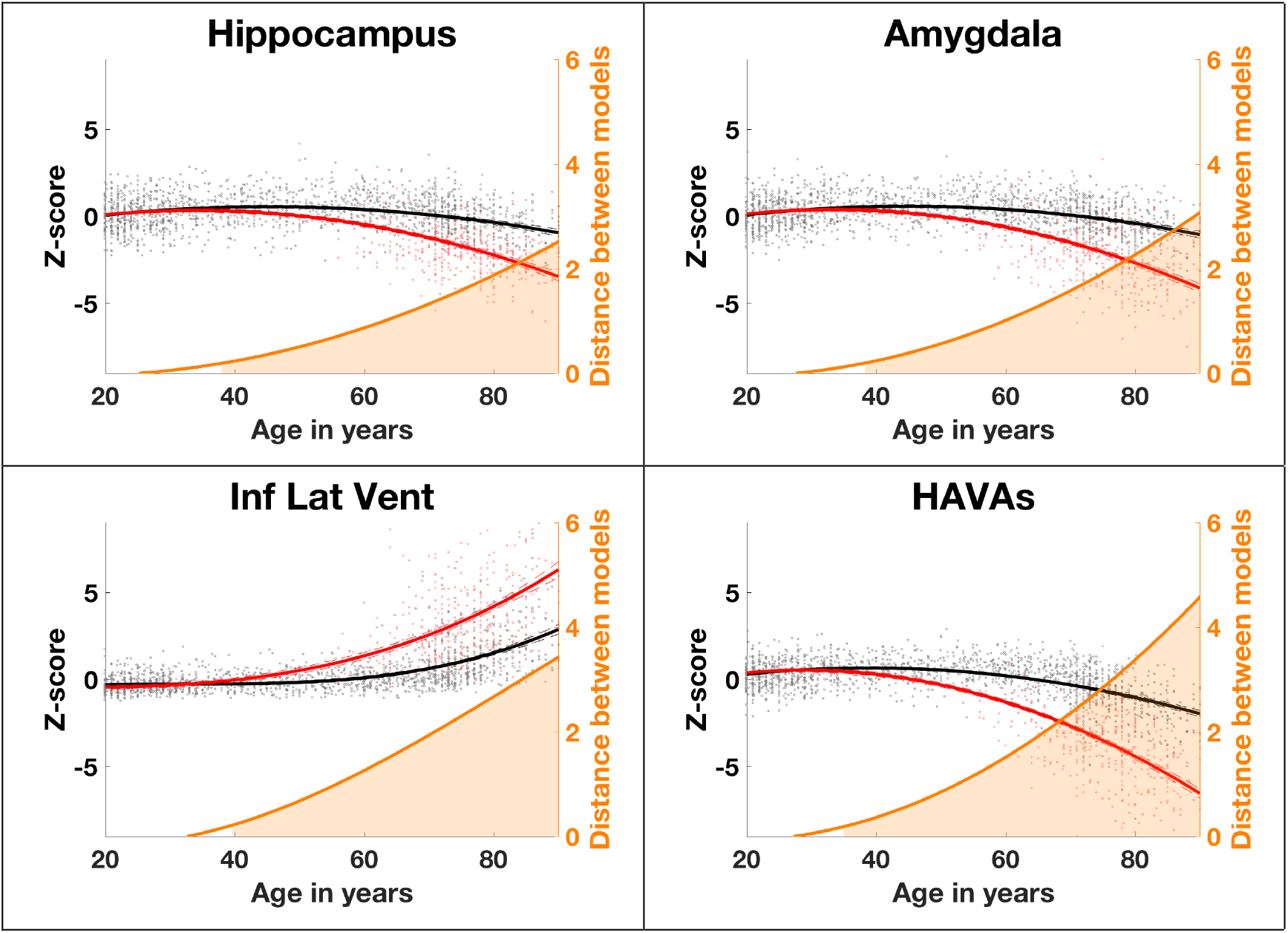
Trajectories based on z-scores of normalized volumes (in % total intracranial volume) for the selected brain structures and the proposed HAVAs for both models (AD in red and CN in black) across the entire lifespan. The prediction bounds of the models are estimated with a confidence level at 95%. The orange curve is the distance between both models in standard deviation. The orange area indicates the time period where confidence intervals of both models do not overlap.

### 3.3 Classification based on multiple lifespan models

To evaluate the classification performance of HAVAs on testing datasets, we performed a comparison with the three most discriminant structures. As shown in Table 4, in all the cases, HAVAs outperformed strategies based on a single structure, in terms of BACC and AUC, demonstrating its higher classification performance. In most of the cases, the second best one was the lifespan model of amygdala that confirmed the results previously obtained in [17]. For diagnostic task (i.e., AD vs. CN), HAVAs obtained 88% of BACC and 84% of AUC on the AIBL database and 89% of BACC and 96% of AUC on the MIRIAD database. Moreover, while developed using only AD and CN subjects, HAVAs obtained 73% of BACC and 78% of AUC for prognosis task (i.e., discriminating between sMCI and pMCI). These results demonstrate the good generalization capabilities of HAVAs on unseen databases and on unseen task.

**Table 4:** Comparison of classification performance of HAVAs compared to individual structures on 3 unseen external datasets (N=1039). The best results are indicated in red and second best in green.

|  | BACC | SPE | SEN | AUC |
| --- | --- | --- | --- | --- |
| **AIBL (467 CN / 82 AD)** |  |  |  |  |
| • HAVAs | **88** | **93** | **83** | **94** |
| • Amygdala | 80 | 85 | 76 | 89 |
| • Hippocampus | 80 | 78 | 82 | 88 |
| • Inferior Lateral Ventricle | 79 | 91 | 67 | 89 |
| **MIRIAD (23 CN / 46 AD)** |  |  |  |  |
| • HAVAs | **89** | **87** | 91 | **96** |
| • Amygdala | 88 | 83 | **93** | 95 |
| • Hippocampus | 74 | 61 | 87 | 87 |
| • Inferior Lateral Ventricle | 86 | **87** | 85 | 91 |
| **ADNI (255 sMCI / 235 pMCI)** |  |  |  |  |
| • HAVAs | **73** | 72 | 74 | **78** |
| • Amygdala | 68 | 69 | 68 | 74 |
| • Hippocampus | 66 | 56 | **77** | 70 |
| • Inferior Lateral Ventricle | 65 | **76** | 54 | 71 |

Figure 3 presents the results of the classification produced by HAVAs on the external datasets. The boundary decision is simply the middle distance between both models. Consequently, false positive are CN subjects (green dots) below orange curve and false negative are AD patients (red dots) above orange curve. Visually, we observed that AD patients exhibited higher variability than CN subjects. Moreover, as expected, most of the MCI were between both models.

**Figure 3:**
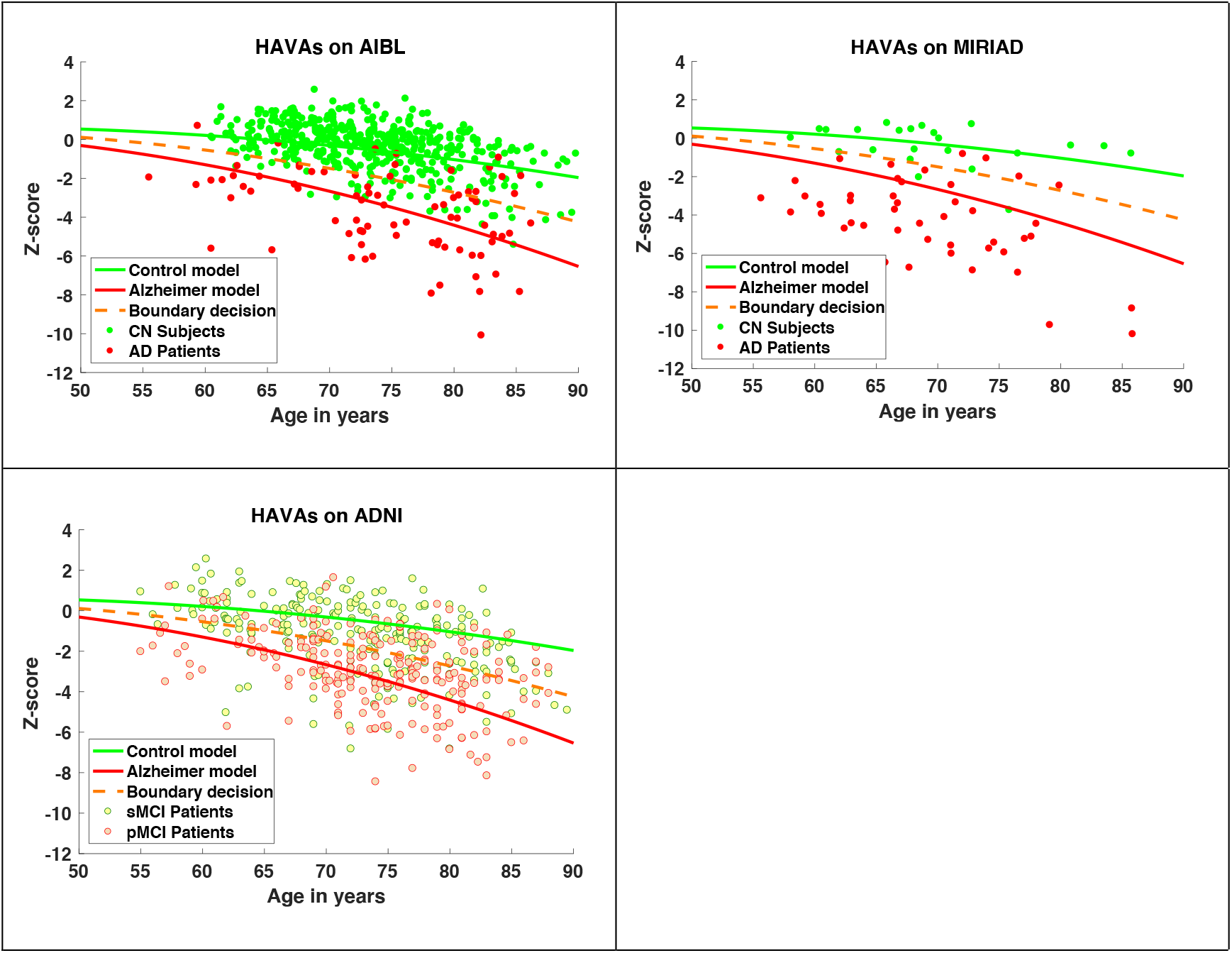
HAVAs classification results on the three external testing datasets. The CN trajectory is in green, the AD trajectory in red and the boundary decision in orange. For AIBL and MIRIAD datasets, CN subjects are in green and AD patients in red. For ADNI dataset, sMCI patients are in yellow and the pMCI patients in orange.

### 3.4 Comparison with state-of-the-art methods

In this section, we compared HAVAs with normative modelling strategy and recent DL methods. As shown in Table 5, HAVAs obtained the best results for both diagnostic and prognostic tasks. Interestingly, the second-best method was the ROI-based CNN involving mostly the same structures as HAVAs. Moreover, the normative HAV model obtained results similar to patch-based CNN and similar prognosis performance compared to all CNN-based methods. In addition, for all the considered structures, the proposed multi-model strategies outperformed single-model based approaches (i.e., normative modelling). Finally, while hippocampus volume is considered a hallmark of AD, normative modelling using hippocampus obtained the worst results with 16% point lower than the proposed multi-model HAVAs.

**Table 5:**
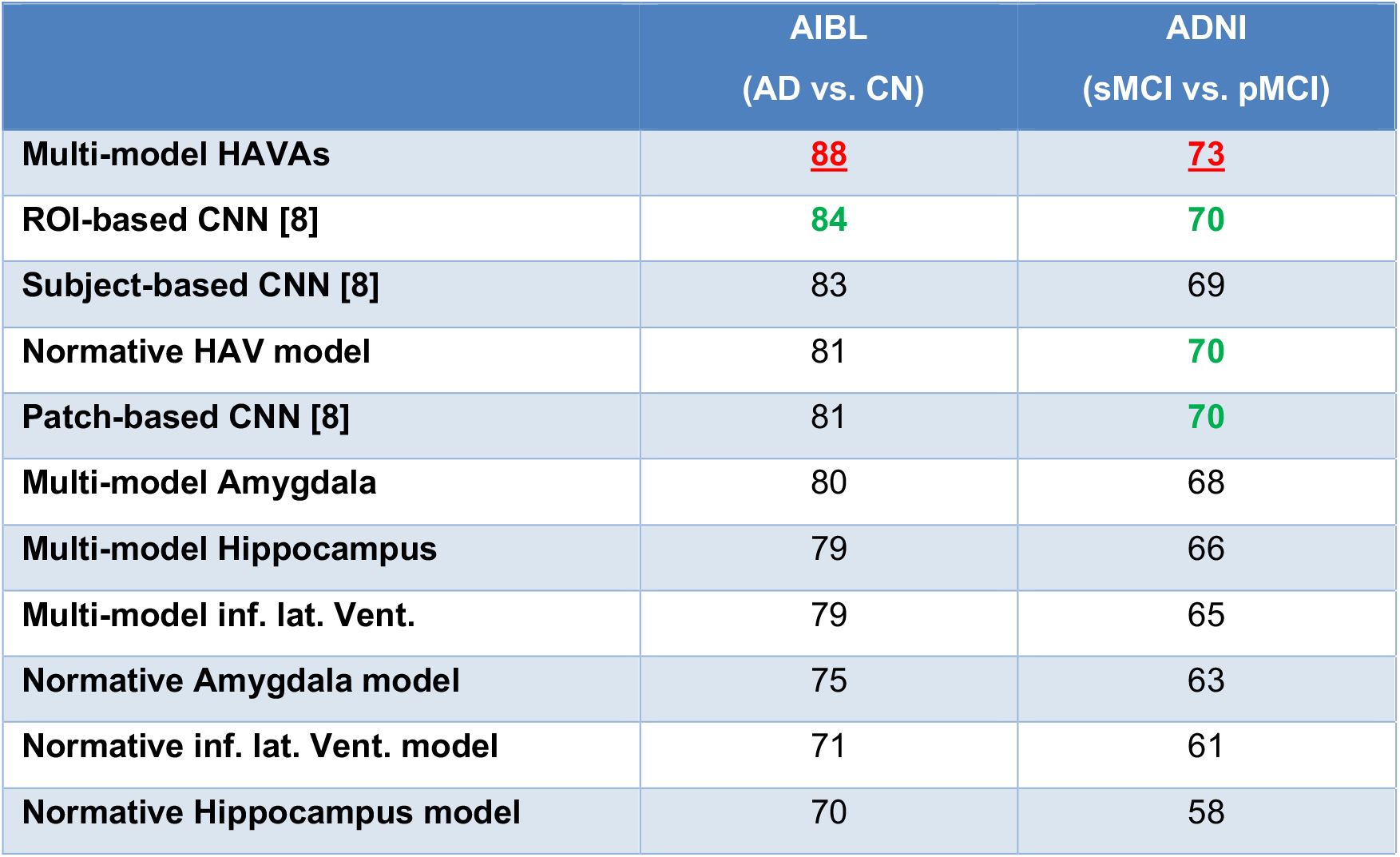
Comparison with state-of-the-art strategies based on normative modelling and recent deep learning methods. BACC is provided for each method for both datasets. For CNN-based methods, the results published in [8] are used. For normative modelling, a threshold of 2σ was used to detect abnormal volumes.

## 4 Discussion

In this paper, we proposed a novel framework for AD detection based on lifespan modelling of the hippocampal-amygdalo-ventricular volume trajectory for both CN and AD. To this end, we first estimated volume trajectories for AD and CN models across the entire lifespan using a large number of subjects. In this study, we analyzed 132 structures, 5 lobes, 4 regional tissues and 3 tissues. This whole brain analysis, in a multi-scale fashion, enabled us to produce a full screening of the diverging brain areas across lifespan between CN and AD. Within the considered brain areas, only 33 showed significantly divergences between AD and CN models. For these 33 brain areas, we estimated the most discriminant lifespan model in terms of classification performance. We found that amygdala, hippocampus and inferior lateral ventricle were the most discriminant structures. These results obtained using AssemblyNet were in line with recent studies based on other segmentation protocols, software or frameworks [17], [28]–[31]. Therefore, we proposed a new AD score based on hippocampal-amygdalo-ventricular volume called HAVAs. This score is based on the distances between the volume of the subject under study and the AD and CN lifespan trajectories. During the validation of HAVAs on three external datasets, we showed that our strategy enables accurate detection of subject having AD, or MCI who will convert to AD in the next 3 years (i.e., pMCI). Finally, we demonstrated the competitive performance of the proposed HAVAs compared to usual normative modelling and recent DL methods.

During our experiments, we showed that models combining several structures (i.e., HAVAs and HAV) outperformed models based on a single structure. This demonstrates the advantage of combining volumes of key structures to improve AD detection. Moreover, our results suggests that methods based on amygdala provide higher accuracy than models based only on hippocampus. The important role of amygdala at the early state of AD has been already observed in the past [17], [30], [32]. Finally, we showed that using several models had beneficial impact for improving classification accuracy compared to single-based model normative approach. We also found that DL methods were in general more accurate than normative modelling approach. Recently, it has been suggested that the combination of both could improve the performance by using normative modelling of learned features [31]. We will investigate this strategy in future works.

To conclude, in addition to improving classification performance, the proposed HAVAs strategy has several advantages over recent DL approaches: - First, HAVAs is conceptually very simple to understand since based on the distance to AD or CN trajectories. This aspect enables an easy interpretability of the results in terms of hippocampal-amygdalo atrophy and concomitant ventricular enlargement. While current DL methods failed to produce relevant explanation on the used features for their decision making [16], HAVAs is fully interpretable and thus is well-suited for clinical practice or pharmaceutical trails. Moreover, the simplicity of HAVAs make it fast and easy to reimplement. A software package including AssemblyNet pipeline and HAVAs estimation will be made freely available as a downloadable Docker^4^ as well as an online pipeline on the volBrain platform^5^ after paper acceptance.

-Second, HAVAs is based on a very low number of parameters and hyperparameters. The use of low order polynomial models for trajectory results in few learnable parameters per trajectory. Thus, using less than ten parameters, HAVAs is able to outperform CNN models involving more than ten million parameters. Moreover, thanks to our volume normalization procedure compensating for inter-subject and inter-structure variabilities, no hyper-parameter is needed to combine hippocampus, amygdala and inferior lateral ventricle volumes. As shown during our experiments, this enables HAVAs to generalize well by being robust to domain shift and efficient on prognosis task.

## Data Availability

All the MRI are freely available through open databases.

## 4.1 Acknowledgements

This work benefited from the support of the project DeepvolBrain of the French National Research Agency (ANR-18-CE45-0013). This study was achieved within the context of the Laboratory of Excellence TRAIL ANR-10-LABX-57 for the BigDataBrain project. Moreover, we thank the Investments for the future Program IdEx Bordeaux (ANR-10-IDEX-03-02, HL-MRI Project), Cluster of excellence CPU and the CNRS/INSERM for the DeepMultiBrain project. This research was also supported by the Spanish PID2020-118608RB-I00 grant from the Ministerio de Ciencia e Innovación.

Moreover, this work is based on multiple samples. We wish to thank all investigators of these projects who collected these datasets and made them freely accessible.

The C-MIND data used in the preparation of this article were obtained from the C-MIND Data Repository (accessed in Feb 2015) created by the C-MIND study of Normal Brain Development. This is a multisite, longitudinal study of typically developing children from ages newborn through young adulthood conducted by Cincinnati Children’s Hospital Medical Center and UCLA and supported by the National Institute of Child Health and Human Development (Contract #s HHSN275200900018C). A listing of the participating sites and a complete listing of the study investigators can be found at https://research.cchmc.org/c-mind. The NDAR data used in the preparation of this manuscript were obtained from the NIH-supported National Database for Autism Research (NDAR). NDAR is a collaborative informatics system created by the National Institutes of Health to provide a national resource to support and accelerate research in autism. The NDAR dataset includes data from the NIH Pediatric MRI Data Repository created by the NIH MRI Study of Normal Brain Development. This is a multisite, longitudinal study of typically developing children from ages newborn through young adulthood conducted by the Brain Development Cooperative Group and supported by the National Institute of Child Health and Human Development, the National Institute on Drug Abuse, the National Institute of Mental Health, and the National Institute of Neurological Disorders and Stroke (Contract #s N01-HD02-3343, N01-MH9-0002, and N01-NS-9-2314, -2315, -2316, -2317, - 2319 and -2320). A listing of the participating sites and a complete listing of the study investigators can be found at http://pediatricmri.nih.gov/nihpd/info/participating_centers.html.

The ADNI data used in the preparation of this manuscript were obtained from the Alzheimer’s Disease Neuroimaging Initiative (ADNI) (National Institutes of Health Grant U01 AG024904). The ADNI is funded by the National Institute on Aging and the National Institute of Biomedical Imaging and Bioengineering and through generous contributions from the following: Abbott, AstraZeneca AB, Bayer Schering Pharma AG, Bristol-Myers Squibb, Eisai Global Clinical Development, Elan Corporation, Genentech, GE Healthcare, GlaxoSmithKline, Innogenetics NV, Johnson & Johnson, Eli Lilly and Co., Medpace, Inc., Merck and Co., Inc., Novartis AG, Pfizer Inc., F. Hoffmann-La Roche, Schering-Plough, Synarc Inc., as well as nonprofit partners, the Alzheimer’s Association and Alzheimer’s Drug Discovery Foundation, with participation from the U.S. Food and Drug Administration. Private sector contributions to the ADNI are facilitated by the Foundation for the National Institutes of Health (www.fnih.org). The grantee organization is the Northern California Institute for Research and Education, and the study was coordinated by the Alzheimer’s Disease Cooperative Study at the University of California, San Diego. ADNI data are disseminated by the Laboratory for NeuroImaging at the University of California, Los Angeles. This research was also supported by NIH grants P30AG010129, K01 AG030514 and the Dana Foundation. The OASIS data used in the preparation of this manuscript were obtained from the OASIS project funded by grants P50 AG05681, P01 AG03991, R01 AG021910, P50 MH071616, U24 RR021382, R01 MH56584. See http://www.oasis-brains.org/ for more details.

The AIBL data used in the preparation of this manuscript were obtained from the AIBL study of ageing funded by the Common-wealth Scientific Industrial Research Organization (CSIRO; a publicly funded government research organization), Science Industry Endowment Fund, National Health and Medical Research Council of Australia (project grant 1011689), Alzheimer’s Association, Alzheimer’s Drug Discovery Foundation, and an anonymous foundation. See www.aibl.csiro.au for further details.

The ICBM data used in the preparation of this manuscript were supported by Human Brain Project grant PO1MHO52176-11 (ICBM, P.I. Dr John Mazziotta) and Canadian Institutes of Health Research grant MOP- 34996.

The IXI data used in the preparation of this manuscript were supported by the U.K. Engineering and Physical Sciences Research Council (EPSRC) GR/S21533/02 - http://www.brain-development.org/.

The ABIDE data used in the preparation of this manuscript were supported by ABIDE funding resources listed at http://fcon_1000.projects.nitrc.org/indi/abide/. ABIDE primary support for the work by Adriana Di Martino was provided by the NIMH (K23MH087770) and the Leon Levy Foundation. Primary support for the work by Michael P. Milham and the INDI team was provided by gifts from Joseph P. Healy and the Stavros Niarchos Foundation to the Child Mind Institute, as well as by an NIMH award to MPM (R03MH096321). http://fcon_1000.projects.nitrc.org/indi/abide/

This manuscript reflects the views of the authors and may not reflect the opinions or views of the database providers.

## Footnotes

1 https://github.com/volBrain/AssemblyNet

2 http://neuromorphometrics.com/Seg/

3 http://neuromorphometrics.com/ParcellationProtocol_2010-04-05.PDF

4 https://github.com/volBrain/AssemblyNetAD

5 http://www.volbrain.net/

